# Attitudes to health promotion among teaching staff in South West England: a qualitative study

**DOI:** 10.1101/2021.08.25.21262589

**Authors:** Jonny Currie

**Author notes:** **Correspondence:** Public Health Wales, 2 Capital Quarter, Cardiff, CF10 4BQ., **E-mail:**. **Authorship** This article has been written by the corresponding author (JC) who wrote the study protocol, conducted interviews and co-led the analysis. Contributions to the study include Dr Rebecca Langford (RL; Senior Research Associate, University of Bristol), Miss Heide Busse (HB; PhD Student, University of Bristol), Mr James Redmore (JR; former Research Assistant, University of Bristol) and Professor Rona Campbell (RC; Professor of Public Health Research, University of Bristol). RL conducted interviews, supported the analysis and commented on the article submission. HB and JR conducted interviews. RC oversaw the study and provided critical insights into the analysis and commented on the article prior to submission.

## Abstract

**Background:** Young people spend much of their formative years in education, making schools appealing environments for health promotion. The World Health Organisation’s Health Promoting Schools framework has been proposed as a useful model. We sought to explore secondary school teachers’ experiences of implementing this model and their attitudes to health promotion.

**Objectives:** To explore teachers’ experiences and perceptions of health promotion and compare practice with the HPS framework for school health.

**Design:** Qualitative study with semi-structured interviews

**Setting:** Nine state comprehensive schools in Bristol and surrounding areas.

**Participants:** 25 teaching staff from school senior leadership teams, those working in health education and other subject teachers.

**Analysis:** Thematic analysis using NVivo 10.

**Results:** Teachers largely described educational approaches with less emphasis on school ethos or environment. Staff supported a role for schools in promoting health but felt restrained by limited family engagement, contradictory school practices, resource constraints and conflicting government policies.

**Conclusions:** Future reforms should ensure health is mainstreamed across school strategies, if we are to create the conditions that promote future generations’ health. Public health must build alliances with educationalists to support the priority-setting of health in school inspections, policy and practice.

**Strengths and limitations of the study:**

- In-depth exploration with teachers from different schools, levels of seniority and with varying experience of health promotion
- Purposive sampling to ensure representation from schools of different geography, Ofsted rating and proportions of students from disadvantaged backgrounds
- Possibility that schools with greater enthusiasm for health promotion disproportionately participated
- Trend towards schools in more affluent areas participating

## Introduction

Young people spend a considerable period of their formative development in education, making them worthy settings for public health promotion [1]. Early initiatives adopted an educational approach focused on providing information to students to encourage healthy lifestyles [2]. Since then, a socio-ecological model has evolved, led by the World Health Organisation (WHO) and its Health Promoting Schools (HPS) framework [3] (see Figure 1). Recognising that educational approaches in health promotion fail to improve health and narrow inequalities, this framework incorporates the school environment and wider community to highlight how schools can become health promoting settings [4]. Schools that promote health according to this framework ensure students have access to quality information on health, learn in an environment which facilitates healthy behavior and mental wellbeing, and develop links with families and the local community for health promotion programmes and community cohesion [3].

**Figure 1.**
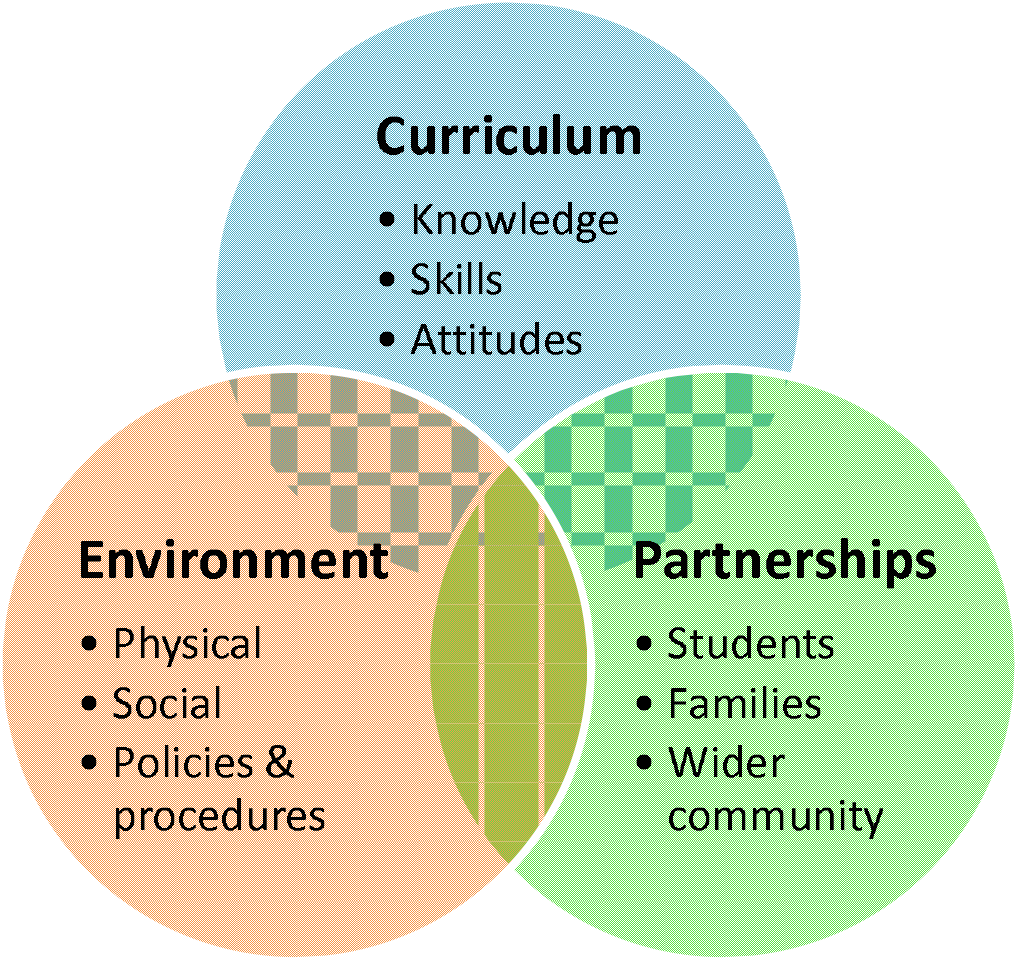
World Health Organisation Health Promoting School (HPS) Framework. Adapted from WHO, 1997 [3].

Recent research has suggested school health promotion to be evidence-based and critical to improving student attainment. A Cochrane review in 2015 concluded the HPS framework to be effective at improving both physical and mental health [5]. Far from distracting from educational outcomes, investment in school health promotion synergistically improves health while improving student attainment [6], [7], [8]. Yet despite this evidence, schools’ capacity in England and Wales to promote health has been significantly constrained of late. A national ‘Healthy Schools’ programme was abolished by government in 2013 [9]. Since 2012 Ofsted no longer includes student wellbeing in its school inspection reports [10]. School autonomy from local authorities has risen significantly after the introduction of academies [11], arguably limiting local authorities influence and reach. Juxtaposed with recommendations that a focus on early years is essential to addressing health inequalities [12], such practice appears contradictory to protecting the health of young people and future generations.

Interestingly, little school health research has engaged with those working in education. One study [13] found teachers largely conceptualised school health within a curricular model. Another found similarly the wider school environment largely neglected in teachers’ discussions [14]. Some authors have found teachers to view health promotion as outside the core business of schools [15], though other studies have found the opposite, provided leadership and policy is supportive [16]. As the professionals who coach young people, teachers’ perspectives are arguably crucial to the development of school health policy. As such, their ‘lived experience’ [17] is vital to understand. This study sought therefore to explore teachers’ perceptions of school health promotion while assessing what relevance the HPS framework on school practice.

## Methods

### Study setting

Bristol is a city in the South West of England with a population of approximately half a million people [18] and borders the local authorities of Bath and North East Somerset and South Gloucestershire. The South West has a large rural population and more favourable rates of employment compared with other English regions, whereas Bristol has areas of significant deprivation, with around 30% being among the 20% most deprived areas in England [19]. Comparisons between Bristol and its surrounding geographies in ethnic diversity demonstrate other significant differences, with the surrounding areas having among the lowest proportions of non-white ethnic groups of all English regions [19], compared with Bristol’s diverse cultural heritage [20].

### Participants

Teaching staff from secondary schools were recruited with purposive sampling from a mixture of urban and rural schools, varying socioeconomic statuses and Ofsted ratings (see Table 1). Inclusion criteria were teaching staff from state comprehensive secondary schools in the Bristol and surrounding area with the ability to provide informed consent. Independent and specialist schools were excluded. Attempts were made to maximise interviewee diversity by recruiting a balance of senior management, those with a responsibility for health and general teaching staff.

**Table 1.** Summary of schools that participated in the study.

| School | Free meal eligibility (%)<sup>1</sup> | Geographical location<sup>2</sup> | Ofsted rating<sup>3</sup> | N. of interviews | Participants |
| --- | --- | --- | --- | --- | --- |
| 1 | 5.8% | Rural | 3 | 2 | PSHE teacher, general teaching staff |
| 2 | 7.5% | Urban | 1 | 3 | Assistant Principal, Senior Learning Co-ordinator, PSHE Co-ordinator |
| 3 | 9.6% | Rural | 1 | 2 | Assistant Head Teacher, PSHE teacher |
| 4 | 5.4% | Rural | 2 | 5 | Head of Year (2), Head of health and social care, Assistant lead of student progress, PSHE Co-ordinator |
| 5 | 11.2% | Urban | 3 | 3 | Head of Year, PE teacher, general teaching staff |
| 6 | 32.4% | Urban | 3 | 3 | Head teacher, Head of Science, general teaching staff |
| 7 | 7.4% | Rural | 1 | 3 | Assistant Head, PSHE/PE teacher, general teaching staff |
| 8 | 8.2% | Urban | 1 | 1 | PSHE co-ordinator |
| 9 | 48.1% | Urban | 2 | 3 | PSHE teachers (2), general teaching staff |

<sup>1</sup> Average free school meal entitlement in UK stated-funded secondary schools for this period was 16.3% [28]
<sup>2</sup> Data from http://www.education.gov.uk/edubase/home.xhtml.
<sup>3</sup> Ofsted ratings: 1 = outstanding; 2 = good; 3 = requires improvement; 4 = inadequate.

### Data collection

Drawing on an interpretivist approach whereby the social realities of others are examined by exploring individual experience and perception [21], semi-structured interviews were conducted using a topic guide [22] based upon themes in the HPS framework and other concepts in school health literature. Participants were encouraged to describe health challenges in their school, inequalities, the impact of health on student attainment and their practice in school health promotion. Study interviews were conducted between December 2012 and March 2013 following a pilot interview and minor amendments to the topic guide. Interviews lasted approximately one hour, were digitally-recorded for transcription and took place on school premises.

### Data analysis

Following transcription of interview recordings by university secretarial staff, data transcripts were inputted to Nvivo 10 [23] for analysis. Interviews were numbered as ‘cases’ and were then coded by identifying key themes and concepts [24] emerging from constant and iterative comparison of transcripts [25]. Transcripts were independently coded by JC and RL with codes emerging deductively using the interview topic guide and inductively through examination of participants’ language. Independent coding of further transcripts was undertaken using a coding framework which demonstrated good inter-rater reliability between interviewers. Authors met repeatedly throughout the analysis period to explore and agree upon emerging themes and make necessary alterations to the code book.

### Ethics

Interviews were conducted with approval at senior management level with full written and informed consent from participants. The study was approved by the University of Bristol Faculty of Medicine and Dentistry Committee for Ethics (FCE application number 111273). All data has been anonymised during analysis to protect participant confidentiality. A copy of the study proposal is available as a supplement.

## Results

### Overview of participants

Twenty-five teaching staff across nine schools participated in the study (see Table 1). Participants came from schools with a range of performance ratings and geographical areas and varied also in specific job role. As demonstrated below, recruitment in schools with higher indices of deprivation proved more challenging.

Staff interviewed gave a variety of comments on their approaches to health promotion in schools, and to factors limiting their success in doing so both inside and outside of the classroom. We outline here some of the revealing points raised by participants, structured under the HPS framework headings of school curriculum, environment and community partnerships.

### School Curriculum

The formal curriculum in the Health Promoting Schools framework was proposed to allow students the opportunity to gain the knowledge, skills and attitudes for healthy lifestyles [3] [5]. This aspect of the HPS framework was the area in which teachers appeared most confident. Teachers cited several health education activities including Personal, Social and Health Education (PSHE), citizenship and tutor lessons. Such education was often delivered within circumscribed areas of the curriculum, by a specific group of teaching staff responsible for health. Interviewees also described numerous initiatives to impart knowledge to students on how they can minimise risky behaviours at an individual level. PSHE specifically covered a range of different health topics aimed at providing young people with the knowledge and skills to make healthy choices in life and to engage them in a debate around healthy lifestyles. Awareness-raising was key to these activities, either within the classroom or more widely in school assemblies:

> *“So, the way I approach it is very sort of educational, so raising awareness about mental health issues, for instances, for both those people who might be experiencing, something and also for everybody else*.*”*
>
> *“Um, the, the stuff discussed in assemblies promotes it, promotes healthy lifestyles, being a, emotionally and physically healthy person on. You know I can think of six or seven assemblies so far this year that I’ve seen that have, you know, really clearly been linked to either healthy eating I can think of three or four that have been linked to healthy eating or perhaps relationships and respect*.*”*

Teachers also cited wider issues in their ability to provide health promotion messages and programmes in schools. Staff confidence was identified as one key constraint with limited training or support being available to empower teachers as agents for health promotion:

> *“For instance, I heard a colleague of mine’s just written a lesson for year ten about first aid. And you, she has got a first aid qualification, she’s looked up the information and then it’s going to be delivered by people. And I think, you know, as a teacher and an adult you are anxious that you don’t say the wrong thing and give the wrong information or misinformation*.*”*

### School Environment

School ethos and environment within the HPS model refers to the potential for school policies, practices and the social and physical environment in a school to positively shape health (WHO, 1997). Comparatively fewer interviewees described health promotion examples of this variety. Where such an approach was discussed, policies that restricted student behaviour were often quoted as examples of ways in which schools could influence the health of their pupils. Several schools for instance took an active stance on energy drinks and mobile phones, usually in the form of an outright ban:

> *“We’ve stopped them having energy drinks. We’ve said that they’re banned from school…they’re allowed to take water into lessons*…*They’re not allowed to bring in like a bottle of pop though*.*”*

Role-modelling was proposed by some interviewees as an opportunity to lead by example: teachers described physical activity as an area in which they could demonstrate a positive lifestyle choice for pupils:

> *“We’ve got our sort of teachers bike shed out the back…I would imagine if we went there now there’d probably be thirty bikes in there…and the pupils are aware that that kind of thing goes on…you know, the pupils are aware of that kind of thing and will talk to members of staff about you know how the, “I went mountain biking at the weekend” or “my colleague went climbing” and things like that so I guess that kind of thing really, you know, sort of promotes it*…*at the end of the day you know, we are role models for them”;*

A small number of teachers cited the importance of open space for physical activity for students’ wellbeing, or of catering companies running a canteen which promoted healthy meal options. For the main part however, few interviewees interviewed explicitly referred to the role of the school environment in relation to health promotion and instead focussed on the opportunities schools had to educate and inform pupils on health and wellbeing issues.

Teachers were able throughout the interview to highlight a wide range of factors that they felt detracted from successful school health promotion. Several participants described contradictory school practices, for instance discouraging physical activity by encouraging students to focus instead on academic work, or rewarding students with unhealthy foodstuffs for good performance:

> *“I think the profile that we allow…physical education to have in schools isn’t high enough, because…if a student needed to do some work, they’d be like oh they can come out of PE lessons do that. And actually I think we’re really trying to come away from that because I don’t think we should be painting the picture that probably of all the subjects they do at school, physical education is actually the thing that they’re most likely to pick up*.*”*
>
> *“[S]o I’ve also had a bit of an issue with prizes always being sweets and chocolates…so that’s bothered me I suppose…It’s done as tutor awards every term for getting sort of, good attendance or sort of winning their tutor challenge. So that’s always for me, been an issue that we reward by giving them sweets. And I don’t feel that that’s a particularly healthy message to give out but other people think differently*.*”*

Several teachers spoke of other limits that had been placed on schools as settings for health promotion. Budgetary cuts were raised by several teachers in the wake of recent local authority funding changes as staff saw previously funded programmes or members of staff no longer in place:

> *“It’s becoming more difficult to involve specialists now, because so many areas have had their education finances cut…sadly where the cuts have come are in these outside agencies that support schools”*

Many spoke of a national focus on academic outcomes and examinations in schools driven by government policy. Several teachers felt unable to prioritise health promotion either in lessons or generally owing to its deprioritised status compared with school exams:

> *“You know I’m being measured on how many of my students have achieved their target minimum grade…I have that in the back of my mind and then I also have, hang on a minute I’m trying to teach these students a lot more because I’m trying to teach them life skills. I’m trying to teach them about healthy eating, nutrition*.*”*

A shared perception was aired that ultimately politics guided priority-setting and the extent of health promotion undertaken in schools. Numerous teachers described recent policy changes that had in their view delegitimised health promotion in favour of other school priorities:

> *“I think the government makes a difference… a change of government has I think deprioritised that part of the curriculum. [U]nder the last government the every child matters policy was a sort of confirmation to head teachers and to local authorities that this really matters and schools will be judged on this as part of OFSTED inspections for example. Whereas I feel that the current government has given the impression that this is less important than it was previously and as long as exam results are good nobody’s going to be too bothered about what kind of health education is provided. [S]chools for necessary reasons jump through hoops which are provided by the government*.*”*

### Partnerships

The Health Promoting Schools philosophy positioned parents and the community as key agents in improving school health, in recognition of their influence on young people’s attitudes, behaviours and social norms [5]. Of those interviewed in our study however parents and the community were described as a more of a barrier to school health promotion. Teachers were aware that home practices had a significant impact on student health and that this impacted on their students’ behaviour:

> *“There’s no routine and structure to their mornings due to the lack of parent involvement and they walk straight into the lesson and you just think ‘oh no it’s just going to go badly today’ you can just see it…their parents haven’t had any control over them or what they’ve been eating and their levels of concentration or motivation [are] not there”*

However, teachers largely voiced frustration regarding families and what was felt to be their unwillingness to collaboratively promote student health. One teacher felt helpless in engaging parents regarding student health:

> *I don’t know what more could be done within school itself to be honest, because I think a lot of it comes with, from parents. And unless the school can find some way of reaching out to parents and persuade them that it’s not a good idea to send little Jonny off with a couple of red bulls and god knows what else. Um, and make sure that parents have given them enough to eat and them to bed on time. I think we should be, I think we should specify in the parent’s guide, year sevens should be bed by nine o’clock and no later at night, you know*.

Some of those interviewed proposed an educational approach to engaging families, proposing outreach to parents and families:

> *“I think home needs to be educated massively. I think that’s where a huge chunk is missing because we, we can do it till we’re blue in the face but when I, when the kids are bringing in their cooking ingredients and you’re looking what their bringing and you go right, OK, hmm. Like you don’t want to be snobbish about but some of the ingredients are shocking and you say well ‘where’s your vegetables?’ Well, ‘My mum never buys vegetables*.*’ “*

Many teachers referred to external agencies in the community who had visited their school to cover specific health topics, including the police, school nurses, theatre companies and sexual health groups. Rather than build relationships with the community to jointly plan health promotion strategies for the school, the community was often viewed more as a ‘resource’ to be utilised in the school setting:

> *“[S]o we get the school nurse comes in to talk each year… on contraception. I organise someone to come in for year seven to talk about the risks of sunbathing and skin cancer…she gives a lecture to them… we sort of try to get some people in because sometimes it’s not very cool for students to take these messages from teachers…But it is from an external person. They seem more happier to believe that they know what they’re talking about”*

## Discussion

This study sought to explore teachers’ attitudes to and experiences of school health promotion in secondary schools in the Bristol and surrounding areas. Participants interviewed endorsed schools taking a proactive stance to student health, though approaches described often comprised a health education model and not the holistic socioecological endorsed by current evidence by the WHO [4] [5]. Participants also expressed a feeling of frustration that policy and education practices were significantly impeding their efforts and appeared under-confident and under-resourced to engage with families and the wider community to tackle the health challenges in their schools.

### Strengths and limitations of the study

Before the results are considered a number of limitations must be explored. Interviews were conducted in one small area and may thus not represent wider practice. The study may have suffered from selection bias with staff with an enthusiasm for health promotion more enthusiastically participating. There was a trend for schools from affluent areas to disproportionally volunteer, though schools in ‘special measures’ would have been less able to participate and may have had greater pressures for academic achievement with resultant negative impacts on student wellbeing. We believe though that our paper has various unique strengths. The qualitative approach adopted allowed a detailed understanding of teachers’ views, providing valuable insights given a relative dearth of such research to date. Participants were diversely spread, from different professional roles and came from schools with varying levels of disadvantage and performance.

### Comparison to wider literature

Several studies have highlighted a predominantly curriculum-focussed or educational approach to school health promotion among teachers [13,14,26]. This study extends these findings by identifying some of the barriers to broader school health interventions at local and national levels. Furthermore, whereas some studies [15] found little appetite for health promotion among teachers, participants in this study believed student health to be part of schools core business. Most research on school health promotion exploring teachers’ attitudes has been conducted in countries outside of the UK, leaving uncertainty of whether the findings apply to the specific political and institutional milieu of schools in England. Given a number of recent large-scale changes to educational policy, school governance through the academies and free schools programme and to the English public health system generally, this study adds a number of critical insights at an opportune time.

### Implications for public health and education policy

As evidence increasingly demonstrates, health and attainment are synergistic agendas [6,7,8], with unequal focus on one in schools limiting the returns an approach that addresses both could provide to the job market and public health. To avoid the diminution of health’s importance in schools and the curriculum, health ought to be given a newly promoted status. The reinstatement of health in Ofsted inspections could for example demonstrate its equal importance to academic outcomes. Prioritising health would avoid health promoting sessions being relegated in favour of academic learning and ensure adequate curricular time is provided to promoting student welfare.

Secondly, public health should advocate immediate health promotion support to equip schools effectively. Without such resources, schools struggle to engage in the wider determinants of student health, evidenced by the lack of environmental approaches throughout interviews. Furthermore, where schools do engage in health promotion, training and resources should highlight the necessity of a socio-ecological approach, given the range of factors which influence students’ health [27]. The reinstatement of the National Healthy Schools Programme which supported a holistic framework to health improvement in schools would for instance remove some of the barriers teachers described to engaging in school health promotion.

Finally, as often the case in public health, it appears school health promotion is as much a political issue as it is a technical one [29]. Since 2010, policy changes to remove health promotion support to schools have stymied teachers’ efforts to promote the health of their pupils [9]. School health promotion encapsulates ambitions society has for future generations. Explicit recognition of this and of the need to engage advocacy should be reflected across school health research, training and practice; proponents of health promoting schools must use their credibility, position and evidence to call for policy changes that better prioritise student health, if the criticisms of the teachers interviewed are to be heeded.

## Conclusion

Teachers appear supportive of an active role for schools in the promotion of young people’s health. Current limits on this enthusiasm require the assertive intervention of public health and policymakers to create the conditions for a comprehensive framework for health promotion to be realised in schools. Future approaches to school health promotion should utilise research, networks and experience to influence policy such that we promote the health of future generations worldwide.

## Supporting information

Study Protocol

## Data Availability

Data from qualitative interviews may be requested from the author.

## Acknowledgements

This research was conducted with the supervision of Professor Rona Campbell and Dr Beki Langford. Dr Liz Anderson kindly provided critical reflections prior to submission.

## Notes

**Significance for public health**: Following a number of significant reforms to education and health policy since 2010, this study explores how schools are engaging in health promotion and the challenges they face in the current political climate. By interviewing teaching staff from secondary schools recruited representatively from areas across the South West of England, the study provides an in-depth perspective of how public health can provide critical insights and support to frontline education services to position schools as health promoting settings. Teachers appear willing participants to the public health agenda, but rightfully ask for the requisite support and leadership from the political system, public health and local authorities, if the health of future generations is to be better prioritised.

**Funding statement** This study was undertaken with the support of The Centre for the Development and Evaluation of Complex Interventions for Public Health Improvement (DECIPHer), a UKCRC Public Health Research Centre of Excellence. Funding from the British Heart Foundation, Cancer Research UK, Economic and Social Research Council [RES-590-28-0005], Medical Research Council, the Welsh Government and the Wellcome Trust [WT087640MA], under the auspices of the UK Clinical Research Collaboration, is gratefully acknowledged.

**Competing interest statement** I declare no conflicts of interests to this submission.

### Competing Interest Statement

The authors have declared no competing interest.

### Funding Statement

This study was undertaken with the support of The Centre for the Development and Evaluation of Complex Interventions for Public Health Improvement (DECIPHer), a UKCRC Public Health Research Centre of Excellence. Funding from the British Heart Foundation, Cancer Research UK, Economic and Social Research Council [RES-590-28-0005], Medical Research Council, the Welsh Government and the Wellcome Trust [WT087640MA], under the auspices of the UK Clinical Research Collaboration, is gratefully acknowledged.

### Author Declarations

University of Bristol Faculty of Medicine and Dentistry Committee for Ethics (FCE application number 111273).

