## Supplementary material for "Attitudes to health promotion among teaching staff in South West England: a qualitative study": Study Protocol

**Perceptions of school health amongst teaching professionals in the Bristol and surrounding area: a qualitative study**

***Study Protocol***

**Sponsor:**

Professor Rona Campbell, School of Social and Community Medicine, 01179287363

**Investigator(s)**:

Dr Jonathan Currie, Research Associate, School of Social and Community Medicine, 0791 756 43 43

Dr Beki Langford, School of Social and Community Medicine, 01179287353

**Protocol details:**

**Location:**

Bristol University School of Social and Community Medicine, Canynge Hall, 39 Whatley Road, Bristol, BS8 2PS

**Rationale:**

Research in recent years has identified a significant influence of childhood and adolescent experiences on adult health and well-being (Galobardes, 2006). Key to this are the behaviours and beliefs shaped during early years which have generated great interest in the value of schools in public health promotion (Langford et al, 2011).

School health programmes traditionally involved the delivery of information or specific health skills to young people in the hope of facilitating healthy lifestyle choices (Lynagh, 1997). Proposals have emerged in recent years however for more comprehensive and integrated programmes that focus on attitudes, behaviours, and the school environment (Deschesnes et al, 2010), or a “settings” approach (St Leger, 2001). A systematic review in 1999 suggested there was “limited but promising” data in favour of health promoting schools (Lister-Sharp, 1999) while a further Cochrane review is currently underway analysing a variety of other more rigorous school health studies (Langford et al, 2011).

The public health community must remain mindful however that schools principal focus is educational outcomes (St Leger, 2001). Child health and educational outcomes may be strongly associated (Powney, 2000) but success of implementation will rely on teachers’ understanding of and preparation in school health promotion (St Leger, 1998).

**Study objectives:**

This qualitative study therefore aims to explore perceptions of Bristol secondary school staff and staff in local authorities and the third sector towards school health promotion and to consider these in the light of current policy for healthy school programmes.

Specifically, the study shall:

(a) Identify health and well-being issues teachers identify of most importance;
(b) Explore the ways in which such health issues are perceived to affect students’ ability to learn; and
(c) Consider necessary resources and support structures to creating a health promoting school.

**Study design:**

This research study will comprise two phases. The first will review current published and grey literature alongside UK government policy on school health promotion to identify key approaches and themes. The second phase will comprise a qualitative study involving semi-structured interviews with teaching staff at state secondary schools in the Bristol and surrounding area and key informants in local authorities and other relevant agencies responsible for education and young people’s health.

This study will recruit teaching professionals from state secondary schools including academies in the Bristol and surrounding area. A snowballing technique shall be used to identify key informants from known individuals in local healthy schools networks. A purposive sampling strategy will be utilised in order to gain insights from key actors in school health including senior management, classroom teachers, personal social and health education staff and members of local authorities or agencies involved in school health.

Participants will be recruited from purposively selected secondary schools in Bristol and surrounding area with representation in rural/urban backgrounds, varying socioeconomic backgrounds determined by proportion of children receiving free meals, varying performance of schools judged by Ofsted and different school sizes.

A total of 10 schools will be targeted with a maximum of 3 teachers from each school although exact numbers shall depend on data saturation during the study. We anticipate an enrolment rate of approximately 40% of teachers contacted from previous study experience. The total study duration shall be six months from recruitment to write-up.

An independent reviewer from the School of Social and Community Medicine shall review the feasibility, methods and aims of the study.

**Methods & analysis:**

Interviews shall be undertaken with participants and shall last approximately hour. The study shall employ the use of an interview guide to structure discussion and provide prompts where appropriate.

Interviews shall explore what participants believe are the key health and well-being issues affecting young people & teachers in schools; what impact health and well-being has on students’ ability to learn; what role they believe schools should adopt in addressing health and well-being issues; and what resources or structures are necessary to creating a health promoting school.

Interviews will be digitally recorded and stored securely on a university database. Digital recordings of interviews shall be fully transcribed. Data will be managed using a framework approach (Smith and Firth 2011). Data coding with NVivo software will be undertaken. A thematic approach to the data analysis will be undertaken with themes and sub-themes and the relationships between them identified. Descriptive and explanatory accounts of the data will be coupled to the data analysis.

**Recruitment:**

Recruitment to the study shall comprise four key steps:

Step 1. Contact with schools through email to the head teacher with a detailed letter explaining the background and purposes of the study

Step 2. Identification of teachers for interviews by head teachers or other member of the senior management team

Step 3. Contact teachers including an invitation letter and information leaflet

Step 4. Identification of key informants through communication with key individuals within local Healthy School networks and local authorities

Schools shall be offered financial reimbursement if necessary for the time staff have taken during interviews out of work or class time such that they can employ a substitute teacher should they wish.

Head Teachers shall receive an invitation letter and information leaflet by post with a reply slip to confirm their interest. Contact will then be made with the school to identify appropriate members of staff who shall receive an invitation letter and information leaflet. Head teachers and staff shall have the opportunity to contact the research team for discussion or clarification if necessary. Key informants from local authorities and agencies involved in school health shall be approached for participation in the study after nomination by our existing contacts and shall receive an invitation letter, information leaflet and consent form to participate.

Interviewees shall give their written consent at the time of interview which shall be at least one week after receipt of the written information materials. All participants have the right to refuse to participate at any point of the study. All participants will be informed they can withdraw at any point without offering a reason.

This study shall not recruit staff from schools for students with specialist educational needs as the health issues relevant to such a school environment are likely to differ significantly from the study question and scope. The study otherwise shall maintain a respect for the diversity of human culture and conditions and therefore shall take full account of ethnicity, gender, disability, age, socio-economic status and sexual orientation in its design, undertaking, and reporting. Individuals will be excluded if their data is incomplete and the information is essential to the study/analysis, or if any reason is given that participating in the study would cause them in any way distress or harm; any exclusion criteria applied will be fully documented in the study write-up.

**Data collection, handling & record keeping:**

Participants will be audio-recorded during interviews. This will be explained in all study communication including the invitation letter, information leaflet and consent form. Data transcription from audio recordings shall be undertaken by a separate transcribing agency.

All personal information arising from the study interviews will be anonymised. Participants will be given a unique identification number and shall not have their name included in the study. Hard copies of data will be stored in a locked filing cabinet. Electronically the data will be stored on a secure database. Digital audio recordings will be stored on a secure, password protected drive. School of Social and Community Medicine policies will also be adhered to regarding handling of data.

The study shall conform in all of its procedures to the Data Protection Act 1998.

**Research governance & ethics:**

The study shall be conducted in compliance with the Research Governance Framework for Health & Social Care and Good Clinical Practice.

A separate reviewer shall assess the procedures and design of the study including its impact value, feasibility, clarity of purpose and suitability of methods.

Ethical approval shall be sought from the University of Bristol Faculty of Medicine & Dentistry Ethics Board.

**Funding:**

Funding for research expenses shall be provided by the DECIHPer centre and shall primarily be used for reimbursing schools for teachers’ time spent in interview and for transcribing fees.

**Outcomes & dissemination of results:**

Data from interviews shall be analysed and included in papers for peer-reviewed journals and/or conference presentations.

Results shall be summarised into a short report and disseminated in an anonymised fashion to participating schools.

The study may be used to inform future research for the DECIPHer centre and may inform the activities of school health networks and agencies.

**Researchers:**

Dr Jonathan Currie is a junior doctor currently placed at the Department of Social and Community Medicine as part of his Academic Foundation Programme.

Dr Beki Langford is a Research Associate and medical anthropologist working at the DECIPHer (Development and Evaluation of Complex Interventions for Public Health Improvement) UKCRC Public Health Research Centre of Excellence in the School of Social and Community Medicine, University of Bristol. She is leading a Cochrane Review on the Health Promoting Schools framework.
